# Covid-19, Lockdowns and Motor Vehicle Collisions: Empirical Evidence from Greece

**DOI:** 10.1101/2020.12.26.20248883

**Authors:** Sotiris Vandoros

**Affiliations:** King’s College London and Harvard T.H. Chan School of Public Health

**Keywords:** Covid-19, lockdowns, motor vehicle collisions, crashes, injury, mortality

## Abstract

Reduced mobility during Covid-19 lockdowns means fewer vehicles at risk of collision, but also an opportunity to speed on empty streets. Other collision risk factors that have changed during the pandemic include alcohol consumption, sleeping patterns, distraction, unemployment and economic uncertainty. Evidence on the impact of the Covid-19 pandemic on motor vehicle collisions is scarce, as such statistics are often released with a delay. The objective of this paper is to examine the impact of the first wave of the pandemic and the first lockdown on motor vehicle collisions and associated injuries and deaths in Greece. Using monthly data at the regional unit level, I provide descriptive evidence and subsequently follow a difference-in-difference econometric approach, comparing trends in 2020 to those of the previous five years while controlling for unemployment and petrol prices. I found a steep decline in collisions, injuries and deaths compared to what would have been otherwise expected. In March and April 2020, there were about 1,226 fewer collisions, 72 fewer deaths, 40 fewer serious injuries and 1,426 fewer minor injuries compared to what would have been expected in the absence of the pandemic.

## 1. Background

Covid-19 has caused over 1.7 million deaths globally. The pandemic has led to a decrease in economic activity and rising unemployment rates.^1^ There have been reports that the pandemic is also affecting health outcomes for people who are not suffering from Covid-19. These may include delayed diagnosis and treatment,^2^ mental health problems,^3^ suicide^4^ and excess mortality in general.^5^

Another outcome that we would expect to be affected by the pandemic and lockdowns is that of motor vehicle collisions (MVC). The mechanisms between conditions associated with the pandemic and collisions are broad, and relate not only to lockdowns, but also to a number of other factors.

First, the most obvious reason is that reduced mobility means fewer cars on the streets being at risk of collision. People’s mobility was reduced as a result of the lockdown, as well as of their own concerns of being exposed to the virus.

Second, lower levels of traffic volume would create the opportunity to speed on empty streets.^6-7^ Speeding is a major collision risk factor, which is also likely to lead to more severe crashes and a higher likelihood of death.

Third, Covid-19 has led to an increase in unemployment, and there is strong empirical evidence than collisions decrease during recessions.^8-9^

Fourth, previous research suggests that financial worries and economic uncertainty are associated with car crashes, possibly due to distraction, frustration and lack of sleep.^10-12^ Inequality and financial issues may affect reduce social cohesion^13^ and make individuals act more selfishly.^14^

Fifth, worrying about loved ones who have Covid-19 may also cause distraction, thus leading to car crashes.^15^

Sixth, sleep length and patterns have been affected by conditions linked to the pandemic.^16-17^

Finally, alcohol consumption increased during the pandemic,^18^ but as bars and restaurants remained closed during lockdowns, this may have taken place mostly at home.

There have been reports of a decrease in collisions and injuries during lockdowns in a number of countries,^19-22^ but a recent study presents evidence of an increase in speeding and deaths in others.^23^ There is also evidence on an increase in speeding in Greece in the early stages of the pandemic.^24^

The objective of this study is to examine whether there was a change in the number of motor vehicle collisions and associated injuries and deaths during the first wave of the Covid-19 pandemic and first lockdown in Greece.

## 2. Data and Methods

I used monthly data on motor vehicle collisions in Greece for the period 2015-2020, obtained from the Greek Statistics Authority. Monthly data included the total number of collisions involving death or injury; the number of deaths; the number of serious injuries; and the number of minor injuries. Data were reported at the regional unit level (there are 51 units in total). Data were extracted on 20 December 2020, and include some preliminary figures that may be adjusted by the data source at a later date. In addition, I obtained data on petrol prices from Eurostat and unemployment rates from the Greek Statistics Authority.

The first Covid-19 case was reported in Greece on 26 February 2020. As a result of increasing infection rates, schools closed on 10 March, followed by closure of bars, cafés and restaurants on 13 March. The first Covid-19 death was reported a few days later, on 12 March, and Greece entered a national lockdown from 23 March to 4 May. As demonstrated in Figure A1 in the Appendix, daily visits to workplaces during the first lockdown were reduced by up to 84%. Mobility relating to retail & recreation was reduced by up to 86%^25^

In order to study the association between the pandemic and collisions, I first used graphs to compare trends in crashes in 2020 to trends in the previous five years. MVCs demonstrate seasonal patterns, which is why we compare trends in 2020 to those in the same months in previous years, rather than between different months in the same year.

However, there are additional factors affecting collisions, including unemployment and petrol prices, so multivariate models are required. Finding a control group is particularly challenging, given that Covid-19 is a global pandemic. In the absence of the pandemic, one would expect that the trends in MVCs would be the same in 2020 as in previous years. Given that MVCs demonstrate seasonal patterns, the difference between crashes in January-February and March-April in 2020 would be similar to the difference between crashes in January-February and March-April in previous years. Therefore, in the empirical model, 2020 is the ‘treatment group’ (in which Covid-19 occurred), and the period from March onwards in every year is the ‘treatment period’ (as that is when the first Covid-19 death occurred in Greece, and when the first lockdown was introduced). A similar approach, using a difference-in-differences econometric model with previous years as a control group, has been followed in previous studies in the absence of a control group involving a different population.^5,26-27^

The difference-in-differences econometric model is presented in Equation 1:

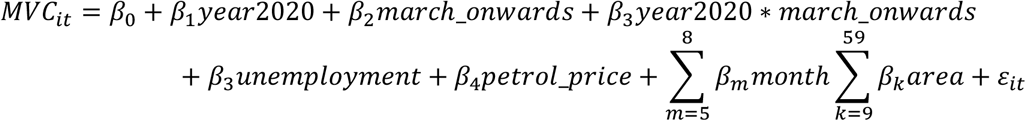

Variable *MVC* represents collision outcomes (number of collisions; deaths; minor injuries; serious injuries). *year2020* is the ‘treatment group’ dummy that takes the value 1 for 2020 and 0 for previous years. *march_onwards* is the ‘treatment period’ dummy that takes the value of 1 for calendar months from march onwards in every year, and zero otherwise. *unemployment* and *petrol_price* represent the monthly unemployment rate and petrol prices, respectively. The model also includes monthly dummies to capture seasonality. Regional unit fixed effects were used to account for heterogeneity across areas, such as population, driving patterns, police presence etc. It is worth noting that population statistics are yet to be released for 2020. Summary statistics are presented in Table A1 in the Appendix.

## 3. Results

Figure 1 shows the trends in MCVs, deaths and minor and serious injuries in years 2015-2020. It is clear from the graphs that there was a steep decline in all four outcomes during the first lockdown (in March and April 2020), compared to trends in previous years. MVC outcomes also remained below the those of previous years’ after the end of the lockdown, although the difference was much smaller than during the lockdown in Spring.

**Figure 1.**
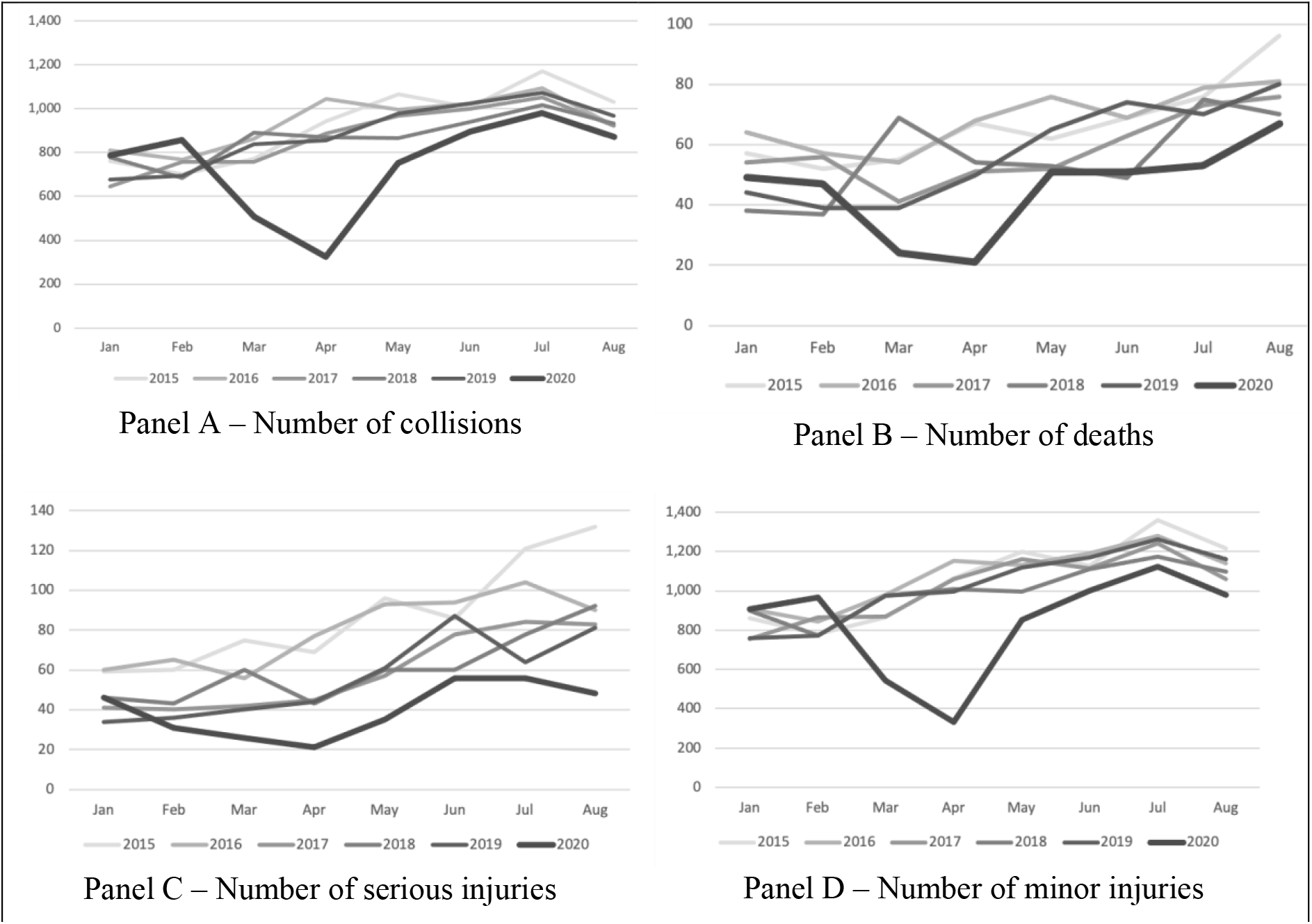
Monthly collisions, deaths, serious injuries and minor injuries in Greece, 2015-2020

MVCs are highly seasonal, so I compared the change between January-February (before the lockdown) and March-April (during the lockdown) 2020 to the corresponding changes in the previous five years (Table 1). While the average increase in MVCs between January-February and March-April in years 2015-2019 was 19.79%, in 2020 there was a *decrease* by 49.49%. Similarly, deaths in 2020 decreased by 53.13% between January-February and March-April, while in the previous five years they had demonstrated a 10% increase.

**Table 1.** Changes in collisions, deaths and injuries per regional unit before and after the first lockdown in Greece in 2020 and the corresponding moths in the previous five years

|  | Jan - Feb | Mar - Apr | % difference |
| --- | --- | --- | --- |
| **Collisions** |  |  |  |
| 2020 | 1,646 | 833 | -49.39% |
| 2015-2019 | 1,455 | 1,743 | 19.79% |
| **Deaths** |  |  |  |
| 2020 | 96 | 45 | -53.13% |
| 2015-2019 | 100 | 110 | 10.00% |
| **Minor injuries** |  |  |  |
| 2020 | 1,874 | 877 | -53.20% |
| 2015-2019 | 1,643 | 1,988 | 21.00% |
| **Serious injuries** |  |  |  |
| 2020 | 77 | 47 | -38.96% |
| 2015-2019 | 97 | 110 | 13.40% |

Data per regional unit (51 units in total)

Results of the difference-in-differences econometric analysis are presented in Table 2. Column 1 provides the results on the total number of collisions. The coefficient of the difference-in-differences interaction term is negative [Diff-in-diff coefficient = −12.020; 95%CI = −18.246 to −5.795; p<0.01], indicating that there were on average 12 fewer monthly collisions per Regional Unit compared to what would have been otherwise expected. This translates to 1,226 fewer collisions for the entire country in total during lockdown months March and April.

**Table 2.** Difference-in-differences regression results, months January – April

| Dependent variable: | (1)<br>Collisions | (2)<br>Deaths | (3)<br>Serious injuries | (4)<br>Minor injuries |
| --- | --- | --- | --- | --- |
| Diff-in-diff interaction term | -12.020***<br>[-18.246 - -5.795] | -0.703***<br>[-1.131 - -0.276] | -0.407**<br>[-0.774 - -0.039] | -13.981***<br>[-21.383 - -6.580] |
| Treatment period dummy<br>(March - April) | 3.706***<br>[1.592 - 5.819] | 0.189*<br>[-0.002 - 0.379] | 0.194*<br>[-0.001 - 0.388] | 4.255***<br>[1.793 - 6.717] |
| Treatment group dummy<br>(2020) | 2.712**<br>[0.340 - 5.085] | 0.216<br>[-0.055 - 0.488] | 0.287**<br>[0.046 - 0.528] | 3.214**<br>[0.039 - 6.389] |
| Unemployment rate | 0.018<br>[-0.168 - 0.204] | 0.032**<br>[0.004 - 0.059] | 0.081***<br>[0.044 - 0.118] | 0.014<br>[-0.214 - 0.241] |
| Petrol prices per litre | -5.043<br>[-15.923 - 5.837] | -0.393<br>[-1.315 - 0.528] | -0.171<br>[-1.202 - 0.861] | -3.308<br>[-15.720 - 9.104] |
| Month dummies | yes | yes | yes | yes |
| Regional Unit Fixed Effects | yes | yes | yes | yes |
| Constant term | 11.395<br>[-7.059 - 29.849] | 0.203<br>[-1.600 - 2.006] | -1.184<br>[-3.386 - 1.018] | 9.940<br>[-11.293 - 31.173] |
| Observations | 1,224 | 1,224 | 1,224 | 1,224 |
| R-squared | 0.965 | 0.780 | 0.796 | 0.961 |
| F-statistic | 71.40 | 13.13 | 14.32 | 62.33 |

Robust ci in brackets. \*\*\* p<0.01, \*\* p<0.05, \* p<0.1

Results on deaths are presented in Column 2. Again, the coefficient of the difference-in-differences interaction term is negative [Diff-in-diff coefficient = −0.703; 95%CI = −1.131 to −0.276; p<0.01]. There were on average 0.703 fewer monthly deaths per Regional Unit than we would have otherwise expected, meaning that during March and April there were 72 fewer deaths in total. Column 3 presents the results on serious injuries [coeff = −0.407; 95%CI = − 0.774 to −0.039; p<0.05] and Column 4 presents results on minor injuries [coeff = −13.981; 95%CI = −21.383 to to 6.580; p<0.01].

In order to study the impact of the pandemic in general rather than just the first lockdown period on MVCs, I extended the study period until August, before the second Covid-19 wave kicked in. Results are presented in Table A2 in the Appendix. Once again, the coefficient of the difference-in-differences interaction term is negative for all four outcomes [Collisions: coeff = −7.294; 95%CI = −10.698 to −3.890; p<0.01. Deaths: coeff = −0.538; 95%CI = −0.874 to −0.203; p<0.01. Serious injuries: coeff = −0.414; 95%CI = −0.752 to −0.077; p<0.05. Minor injuries: coeff = −8.878; 95%CI = −13.123 to −4.633; p<0.01]. However, the relative reduction in MVC outcomes over the entire period is on average smaller than when considering only the first two months of the pandemic.

## 4. Discussion

This study examined the impact of Covid-19 on MVC in Greece during the first wave of the pandemic. I found that during March and April 2020, there were about 1,226 fewer collisions (62% reduction), 72 fewer deaths (68% reduction), 40 fewer serious injuries (48%) and 1,426 fewer minor injuries (63%) compared to what would have been expected in the absence of the pandemic. Extending the study period until August suggests that during the entire study period there was a total reduction by 2,232 collisions, 165 deaths, 127 serious injuries, and 2,716 minor injuries.

Results of this study are particular to the first wave of the pandemic in Greece, which saw a relatively small number of coronavirus cases and deaths, and there was little evidence of effects on other outcomes (e.g. there was no reported increase in suicides.^28^ Results are not necessarily generalisable to other countries that have different types of restrictions (if any) or the second wave in Greece, where the number of cases, hospitalisations and deaths was much larger than the first wave.

There are a number of reasons behind any change in the number of MVC during the pandemic. These include factors that affect traffic volume, i.e. lockdowns, fear of catching Covid-19 and unemployment; and factors that affect driving behaviour, i.e. speeding;^6^ distraction;^15^ uncertainty;^11^ alcohol;^18^ and change in sleep duration, quality and patterns.^16^ The multivariate nature of the relationship between Covid-19 and MVC makes disentangling the effects of individual factors challenging.

The findings of this paper add to the existing literature that found a change in MVC, injuries and deaths during the pandemic, whether they report an increase^19-22^ or a decrease.^23^ The findings also relate to the discussion on excess mortality and other spillover effects^5,29^ and provide evidence on yet another possible factor contributing to the impact of Covid-19 on other health outcomes.

Overall, the Covid-19 pandemic has cost many lives, stretched heath system capacity, led to recessions and unemployment and has caused many other health problems. The reduced number of collisions and related fatalities are just a small positive spillover effect, which is much smaller than the overall negative effects of the virus, and also came as a result of a further negative consequence of the pandemic: that of reduced mobility and freedom.

## Data Availability

The data used on collisions and unemployment are freely available on the website of the Greek Statistics Authority. Petrol prices are provided by Eurostat and are freely available online.

## Ethics approval

The data used were aggregate anonymous data, so ethics approval was not required.

## Checklist

There is no relevant checklist of observational studies.

## Appendix

**Figure A1.**
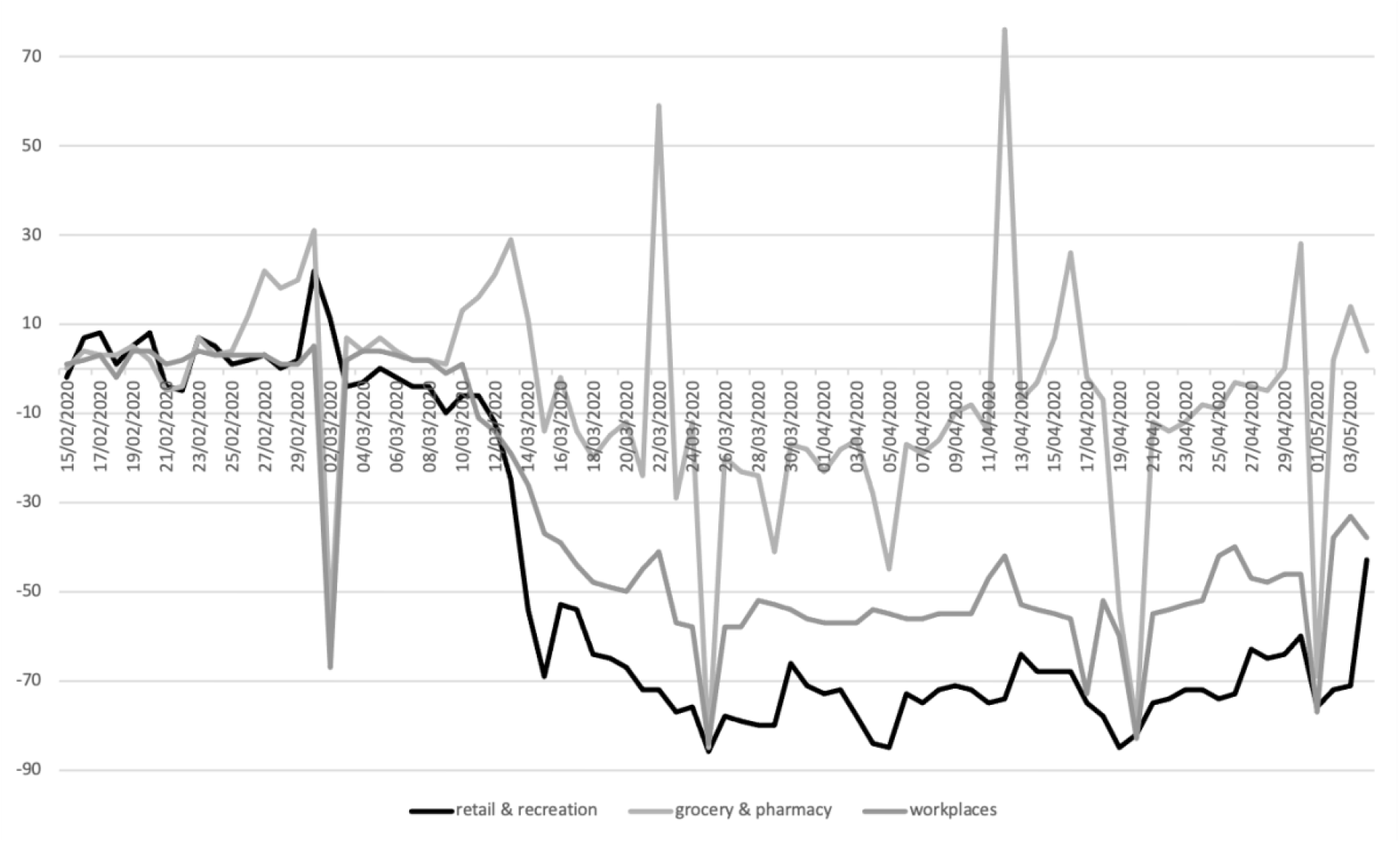
Mobility trends before and during the first lockdown in Greece. Based on data from Google COVID-19 Community Mobility Reports, 2020

**Table A1.** Summary Statistics

| Variable | Mean | Std. Dev. | Min | Max |
| --- | --- | --- | --- | --- |
| Collisions | 17.21 | 64.62 | 0 | 572 |
| Deaths | 1.15 | 2.40 | 0 | 23 |
| Serious injuries | 1.27 | 2.78 | 0 | 28 |
| Minor injuries | 19.36 | 73.51 | 0 | 634 |
| Treatment period dummy (March - April) | 0.75 | 0.43 | 0 | 1 |
| Treatment group dummy (2020) | 0.17 | 0.37 | 0 | 1 |
| Unemployment rate | 20.77 | 3.40 | 14.9 | 27.4 |
| Petrol price per litre in Euros | 1.50 | 0.09 | 1.32 | 1.65 |

*N*=2,448. Data at regional unit level.

**Table A2.** Difference-in-differences regression results, months January – August

| Dependent variable: | (1)<br>Collisions | (2)<br>Deaths | (3)<br>Serious injuries | (4)<br>Minor injuries |
| --- | --- | --- | --- | --- |
| Diff-in-diff interaction term | -7.294***<br>[-10.698 - -3.890] | -0.538***<br>[-0.874 - -0.203] | -0.414**<br>[-0.752 - -0.077] | -8.878***<br>[-13.123 - -4.633] |
| Treatment period dummy<br>(Mrch - August) | 5.549***<br>[3.613 - 7.485] | 0.709***<br>[0.514 - 0.905] | 1.032***<br>[0.782 - 1.281] | 6.952***<br>[4.675 - 9.228] |
| Treatment group dummy<br>(2020) | 3.161***<br>[1.113 - 5.210] | 0.186<br>[-0.082 - 0.454] | 0.256**<br>[0.005 - 0.507] | 4.028***<br>[1.338 - 6.718] |
| unemployment rate | 0.167*<br>[-0.030 - 0.364] | 0.024**<br>[0.002 - 0.046] | 0.094***<br>[0.064 - 0.124] | 0.185<br>[-0.051 - 0.421] |
| petrol prices per litre | -1.805<br>[-9.266 - 5.655] | -0.504<br>[-1.201 - 0.193] | 0.613<br>[-0.280 - 1.506] | -1.635<br>[-10.510 - 7.240] |
| month dummies | yes | yes | yes | yes |
| Regional Unit Fixed Effects | yes | yes | yes | yes |
| Constant term | 2.658<br>[-11.461 - 16.777] | 0.455<br>[-0.945 - 1.856] | -2.725***<br>[-4.600 - -0.851] | 2.622<br>[-14.305 - 19.549] |
| Observations | 2,448 | 2,448 | 2,448 | 2,448 |
| R-squared | 0.969 | 0.773 | 0.758 | 0.965 |
| F-statistic | 117.3 | 24.43 | 24.55 | 96.95 |

Robust CI in brackets. \*\*\* $p < 0.01$ , \*\* $p < 0.05$ , \* $p < 0.1$

